# Stakeholder Interest, Power and Operational Capacity Analysis in the Prevention and Control of Genital Chlamydia in Youths 15–24 Years Old in Mali and Burkina Faso

**DOI:** 10.64898/2026.03.09.26347931

**Authors:** Modibo Sangaré, Moussa Niare, Dramane Sogodogo, Fatoumata Doumbia, Kassoum Alou Ndiaye, Bekaye Coulibaly, Karim Traore, Seidina A Diakite, Samba Diarra, Romain Dena, Mariam Toure, Bourama Keita, Abdrahamane Anne, Emile Badiel, Daouda S Niare, Adama Ouedraogo, Désiré Tassembedo, Mamady Traore, Madina Konaté, Mamadou Diop, Daouda Fomba, Oumar Sidibé, Modibo Kouyate, Aissatou Soumana Billo, Privat Ogniwa, Christelle Fatondji, Ichiaka Moumine Koné, Ousmane Traore, Hama Diallo, Boulaye Sanogo, Ousmane Maiga, Mamadou Dolo, Boureima Guindo, Housseini Dolo, Sory Ibrahim Diawara, Souleymane Dama, Sekou Bah, Kassoum Kayentao, Fatoumata Traore, Cheick AT Traore, Kalifa Keita, Fumiko Shibuya, Jun Kobayashi, Georgina Yeboah, Mahamadou Diakite, Akhere A Omonkhua

**Affiliations:** Faculty of Medicine and Odontostomatology (FMOS), USTTB, Bamako, Mali; Executive Secretariat of the High National Council for the Fight against AIDS (SEHCNLS), Bamako, Mali; Faculty of Pharmacy (FAPH), USTTB, Bamako, Mali; Association Malian Network for Health, Research, Education and Development (MNREHD Association), Bamako, Mali; African Institute of Public Health (IASP), Ouagadougou, Burkina Faso; Permanent Secretariat of the National Council for the Fight against AIDS and STIs (SP/CNLS-IST), Ouagadougou, Burkina Faso; Health Sector Programme for the Fight against AIDS and Sexually Transmitted Infections and Viral Hepatitis (PSSLS-IST/HV), Ouagadougou, Burkina Faso; Sectoral Unit for the Fight against AIDS, Tuberculosis and Viral Hepatitis (CSLS-TBH), Ministry of Health and Social Development (MSDS), Bamako, Mali; Centre of Excellence in Bioinformatics (ACE-B), USTTB, Bamako, Mali; Samu Social, Niamey, Niger; Savè-ouèssè Zone Hospital, Parakou, Benin; National Institute of Public Health (INSP), Bamako, Mali. BP: 1771; NGO Enda Mali, Bamako, Mali; Directorate General of Health and Public Hygiene (DGSHP), Bamako, Mali; Department of Global Health, Graduate School of Health Sciences, University of the Ryukyus, 1076 Kiyuna, Ginowan, Okinawa, 901-2725, Japan; African Forum for Research and Education in Health (AFREhealth), Kumasi, Ghana; University of Sciences, Techniques and Technologies of Bamako (USTTB), Bamako, Mali; Africa Research Excellence Fund (AREF), Medical Research Council Unit, The Gambia @ London School of Hygiene and Tropical Medicine, Faraja, Banjul, The Gambia

**Keywords:** Stakeholder, Interest, power/influence, capacity, chlamydia, youths aged 15-24, Mali, Burkina Faso

## Abstract

**Introduction**: *Chlamydia trachomatis* infection is very common in young people aged 15-24 years old in Sub Saharan Africa. **Aim**: This study aimed to identify key actors involved in the prevention and control of genital chlamydia among young people aged 15–24 and to analyze their positioning and level of commitment to ensure the sustainability and scale-up of the Pfizer Chlamydia Project led by the Faculty of Pharmacy (FAPH). **Methods:** A quantitative stakeholder analysis was conducted from April 2025 to February 2026, with data collection between September 18, 2025 and January 27, 2026. Stakeholders were evaluated using a standardized grid based on three dimensions: interest (0–20), power/influence (0–12), and operational capacity (0–12). **Results:** We found high levels of stakeholder interest (13.0–19.0), with RJASR/PF and national coordination structures demonstrating the strongest engagement. Power/influence scores varied widely (5.0–11.5), with national institutions exerting the greatest influence. Operational capacity scores were relatively homogeneous (8.0–11.0). Coefficients of variation indicated low to moderate agreement for interest and operational capacity, but higher variability for power/influence, particularly among community organizations. **Discussion**: Our findings highlight key partners for strengthening and sustaining chlamydia prevention initiatives in Mali and Burkina Faso. The stakeholder analysis highlights a complementary institutional and community system supporting genital chlamydia prevention among youth in Mali and Burkina Faso. National coordination bodies demonstrated high interest, influence, and operational capacity, confirming their central role in strategic governance and resource mobilization. Technical structures also showed strong capacity for implementing surveillance and screening activities. In contrast, community-based organizations exhibited strong engagement and operational presence but more limited strategic influence, reflecting their field-oriented role in reaching vulnerable youth. Variability in power assessments suggests differing perceptions of institutional influence. Overall, effective project sustainability depends on strengthened collaboration, inclusive governance, and capacity-building to better integrate community actors into decision-making processes.

## 1. Introduction

The sexual and reproductive health of adolescents and young people is a major public health issue in low- and middle-income countries, particularly in sub-Saharan Africa. This age group, which represents a significant proportion of the population, faces multiple vulnerabilities, including insufficient sex education, restrictive socio-cultural norms, limited access to appropriate health services, and persistent stigma around sexuality (Hu et al., 2025; Michalow et al., 2025; Sameni et al., 2025).

In this context, sexually transmitted infections (STIs) are a public health problem of particular concern among young people aged 15 to 24. Among these, genital chlamydia, caused by *Chlamydia trachomatis*, is one of the most common bacterial STIs. Its prevalence is estimated to be about 4.2% in women and 2.7% in men (WHO, 2022). The predominantly asymptomatic nature of the infection promotes its silent transmission and delays diagnosis, exposing young women in particular to severe complications such as pelvic inflammatory disease, infertility, and ectopic pregnancies (WHO, 2022).

In sub-Saharan Africa, the fight against bacterial STIs among adolescents and young people often remains relegated to the background, overshadowed by the priorities given to the prevention and management of HIV/AIDS. Yet, although less lethal, STIs such as genital chlamydia have major health and social consequences, constituting a major cause of infertility among young couples (Rowley et al., 2019).

In Mali and Burkina Faso, the available data suggest a significant, although probably underestimated, circulation of bacterial STIs among young people aged 15 to 24 years. This situation is exacerbated by many barriers, including structural barriers such as the lack of youth-friendly health services, socio-cultural constraints related to taboos surrounding sexuality, socio-economic difficulties (cost of care, limited mobility), as well as organizational constraints, including the low integration of routine STI testing into existing health services (UNFPA, 2021).

It is in this context that the Pfizer Chlamydia Project was initiated, aimed at strengthening the prevention, screening, and management of genital chlamydia in young people aged 15 to 24 in Mali, Burkina Faso, Niger, and Benin. The implementation of this project is based on the mobilization of a variety of actors, including national institutions, technical structures, community-based non-governmental organizations, technical and financial partners as well as international organizations. However, the sustainability and scale of a public health response depend largely on the level of commitment, influence, and operational capacity of these stakeholders. In this respect, the Interest-Power-Operational Capacity analysis is a relevant strategic tool for understanding the dynamics of actors and effectively orienting the governance of the project.

Thus, the objective of this study was, on the one hand, to identify the key actors involved in the prevention and control of genital chlamydia in young people aged 15 to 24, as well as the operational relays and levers of action available, and, on the other hand, to analyze their positioning and level of commitment in order to ensure the sustainability and scaling up of the Pfizer Chlamydia Project led by the Faculty of Pharmacy (FAPH).

## 2. METHODOLOGY

### 2.1. Scope of the study

This study was conducted in Mali and Burkina Faso as part of the Pfizer Chlamydia Project led by the Faculty of Pharmacy (FAPH), University of Sciences, Techniques and Technologies of Bamako (USTTB), Bamako, Mali from January 2025 to March 2026, the objective of which is to develop educational materials to strengthen the prevention, screening, and management of genital chlamydia in young people aged 15 to 24 in Mali, Burkina Faso, Niger, and Benin. The present study was more specifically part of the process of developing and strategically orienting the interventions of the project, in particular through the identification and analysis of stakeholders involved in the prevention and control of genital chlamydia, with a view to strengthening governance, sustainability, and scale-up of actions.

### 2.2. Type and Period of the Study

This was a quantitative analysis based on the attribution of scores to the different dimensions analysed - namely interest, power/influence, and operational capacity. This study ran from April 2025 to February 2026, with a specific period of data collection and analysis between September 18, 2025 and January 27, 2026.

### 2.3. Study population

The study population consisted of sexual and reproductive health program managers, representatives of public institutions, national and international non-governmental organizations (NGOs), as well as youth associations involved in the prevention and management of STIs in Mali and Burkina Faso.

#### 2.3.1. Inclusion criteria

Participants who met the following criteria were included in the study:

◦ Be in charge or manager of a sexual and reproductive health program or member of an NGO/association working in the field of sexual and reproductive health in Mali and Burkina Faso;
◦ Have voluntarily agreed to participate in the study and signed an informed consent.

#### 2.3.2. Non-inclusion criteria

Not included:

◦ individuals/organizations that have no connection with sexual and reproductive health or STI prevention;
◦ s that consented but did not provide their responses before the data were analysed.

#### 2.3.3. Sampling Method

Interest-based sampling was used. At least eight (8) stakeholders per country have been targeted, i.e. 16 in total (Table 1a), comprising at least three (3) public structures, two (2) NGOs, two (2) associations and one (1) network or federation of associations of people living with chronic STIs, including HIV. The minimum sample size was not applicable.

**Table 1:**
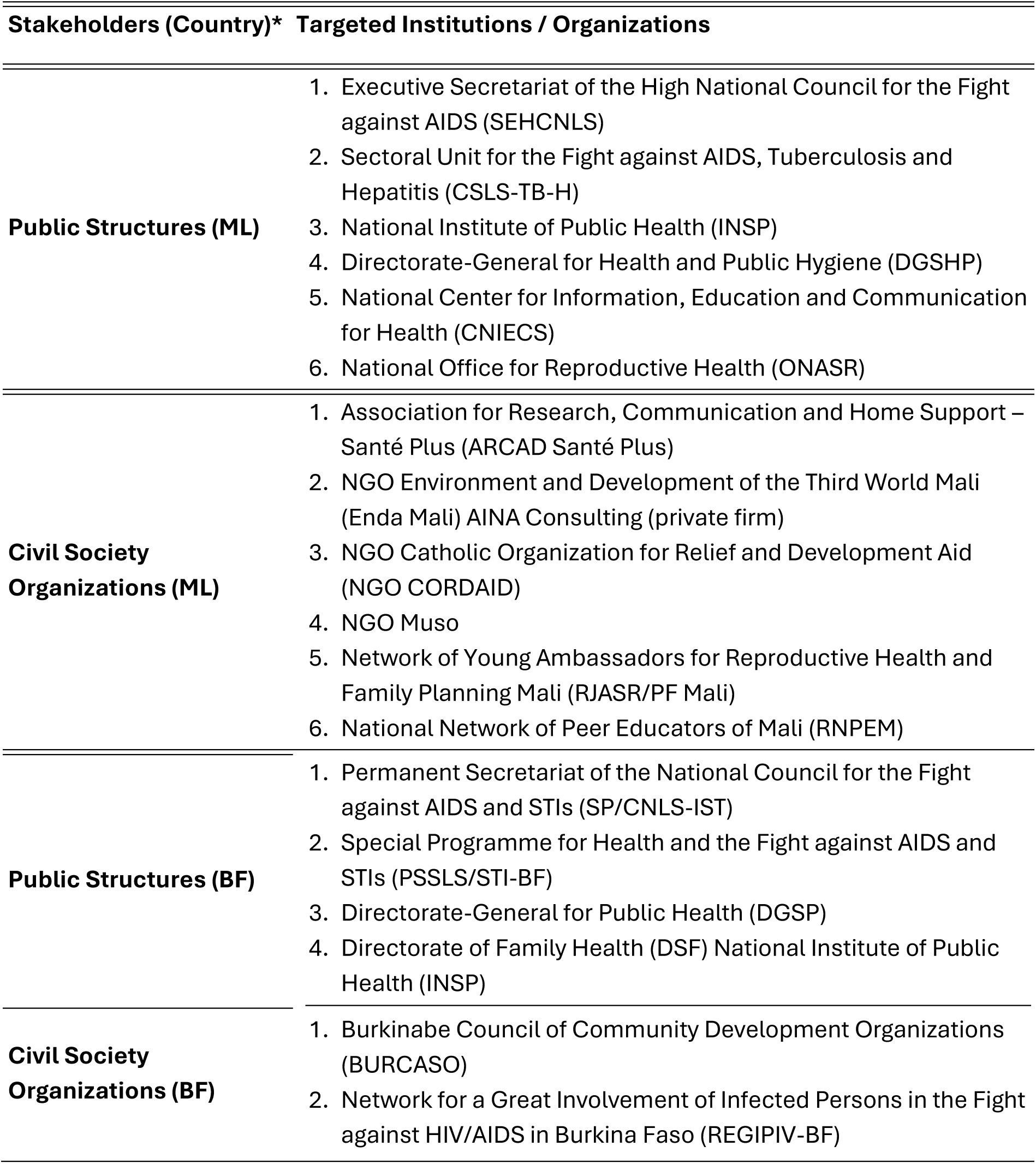
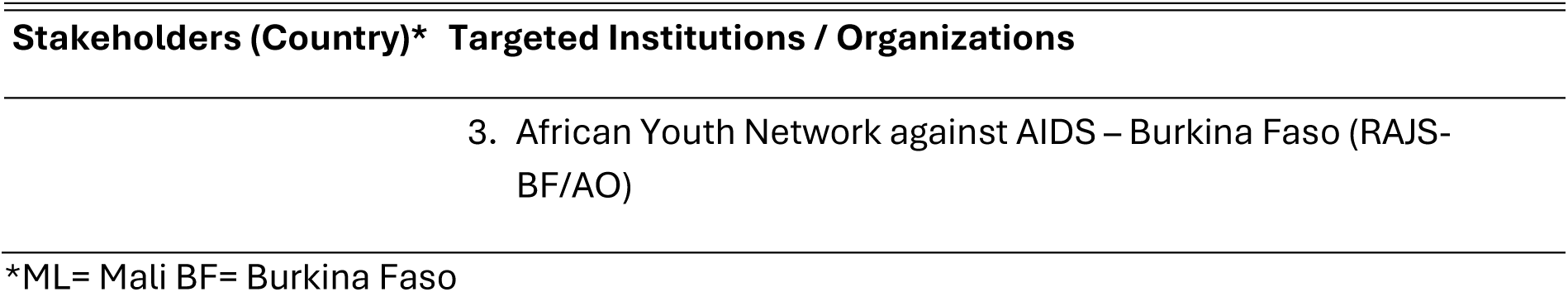
List of targeted stakeholders in Mali and Burkina Faso.

**Table 1b:** Categorization of Stakeholders by Status and Potential Role in the Project in Mali and Burkina Faso.

| **No.** | **Stakeholders** | **Country** | **Type of actors** | **Role Potential in the Project** |
| --- | --- | --- | --- | --- |
| 1 | SE/HCNLS | ML | National Institution | HIV/STI strategic management |
| 2 | SP/CNLS-IST | BF | National Institution | National HIV/STI coordination |
| 3 | CSLS-TBH | ML | Technical structure | Laboratory monitoring and support |
| 4 | PSSLS-STI/HV | BF | Technical structure | Technical support and supervision |
| 5 | NGO ENDA Mali | ML | NGO | Community Outreach and Mobilization |
| 6 | NGO Muso | ML | NGO | Prevention and management |
| 7 | BURCASO | BF | NGO | Prevention and community testing |
| 8 | REGIPIV-BF | BF | NGO | Implementation in the field of STIs/HIV |
| 9 | RNPEM | ML | Network/Association | Youth Peer Education |
| 10 | RJASR/PF Mali | ML | Network/Association | Awareness-raising and youth relays |

\*ML= Mali BF= Burkina Faso NGO ENDA Mali: the Malian national branch of the Enda Third World network BURCASO: Burkinabe Council of Community Development Organizations REGIPIV-BF: Network for a Greater Involvement of People Infected and Affected by HIV/HIV-AIDS RJASR/PF: Network of Young Ambassadors for Reproductive Health/Family Planning RNPEM: National Network of Peer Educators of Mali PSSLS-IST/HV: Health Sector Programme for the Fight against AIDS and Sexually Transmitted Infections and Viral Hepatitis SP/CNLS-IST: Permanent Secretariat of the National Council for the Fight against AIDS and STIs CSLS-TBH: Sectoral Unit for the Fight against HIV/AIDS, Tuberculosis and Viral Hepatitis SE/HCNLS: Executive Secretariat of the National High Council for the Fight against AIDS

**Table 1c:**
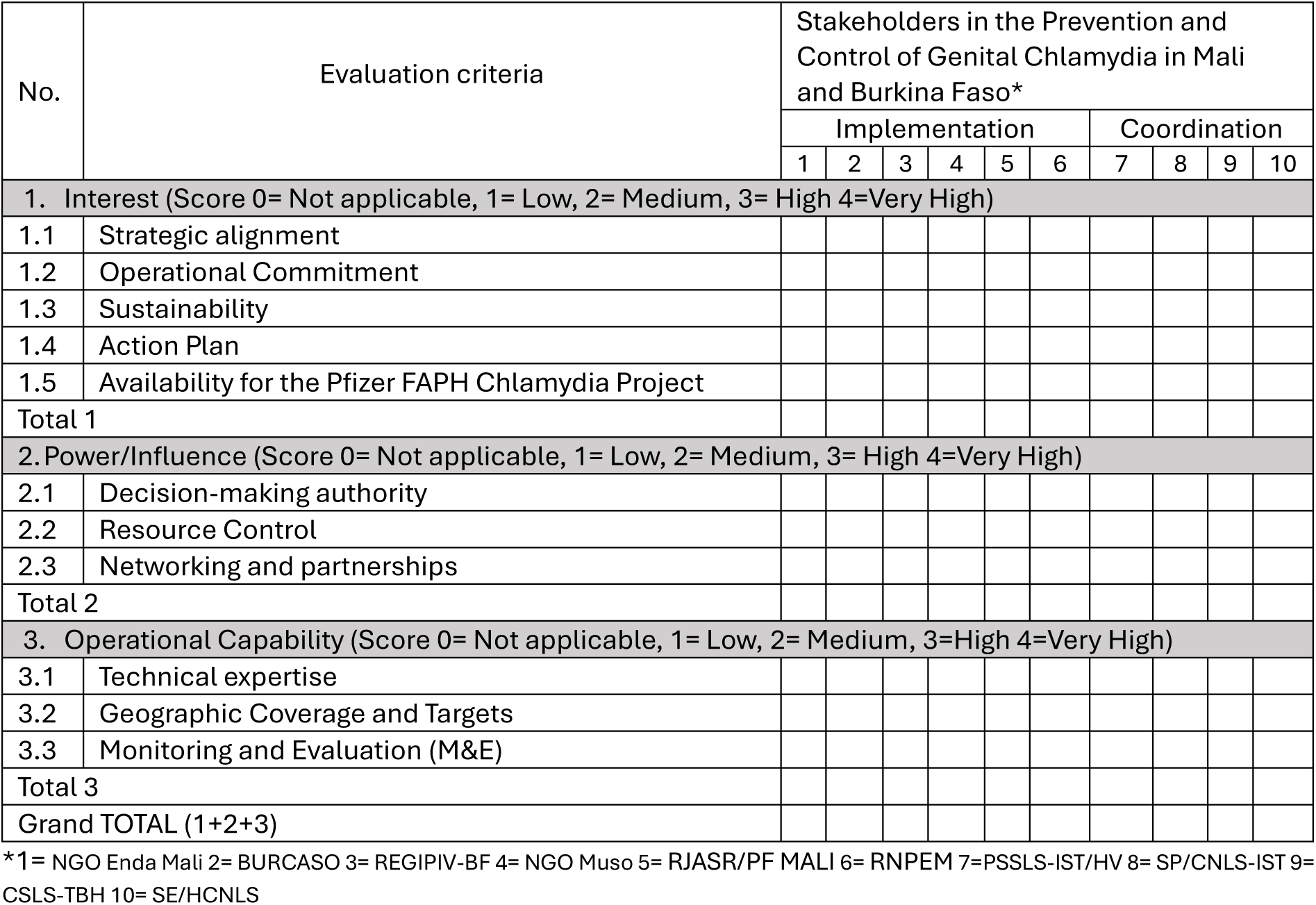
Evaluation Grid of the Interests, Power/Influence and Operational Capacity of the Stakeholders of the Pfizer HPA Chlamydia Project.

The selection of stakeholders was based on the availability of participants, their strategic role and their level of involvement in the sexual and reproductive health of young people in general and the prevention and control of STIs in particular among young people aged 15-24 years.

### 2.4. Conduct of the investigation

### 2.5. Data Collection and Analysis

#### 2.5.1. Data Collection

A self-administered 24-item electronic questionnaire (see Annexes), developed in Microsoft Word 2019, was sent by email to the first leaders of the targeted stakeholders in Mali and Burkina Faso. They completed the questionnaire independently and returned it by email during the data collection period. The responses were then collected, verified, and compiled into a single Microsoft Word 2019 document for analysis.

The information collected was summarized in the form of 1-page summary sheets by each of the ten (10) stakeholders (**Table 1b**), intended for the five (5) independent evaluators.

The evaluation was based on a standardized grid articulated (**Table 1c**) around three (3) main dimensions:

◦ Interest (score 0–20): level of commitment, buy-in and motivation to support the project;
◦ Power/Influence (score 0–12): ability to influence decisions, strategic directions, and sustainability of the project;
◦ Operational capacity (score 0–12): available human, technical, logistical, and organisational resources.

Each rater independently assigned a score for each dimension and for each stakeholder. The completed grids were then consolidated into a single table for the calculation of total scores and the categorization of the different dimensions (**Table 2a - Table 2c**).

**Table 2a:**
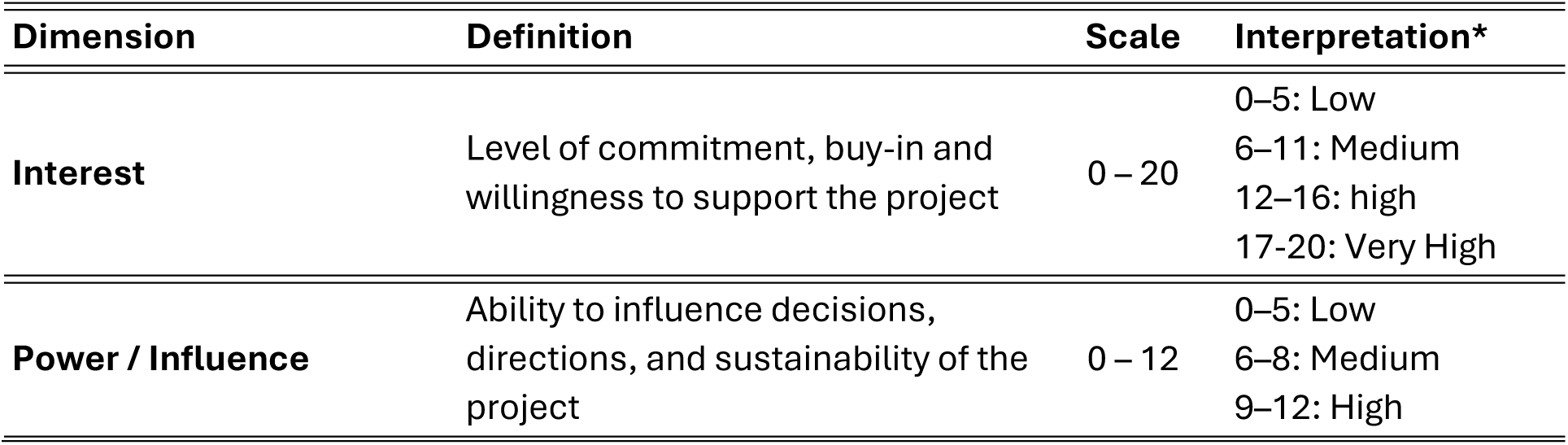
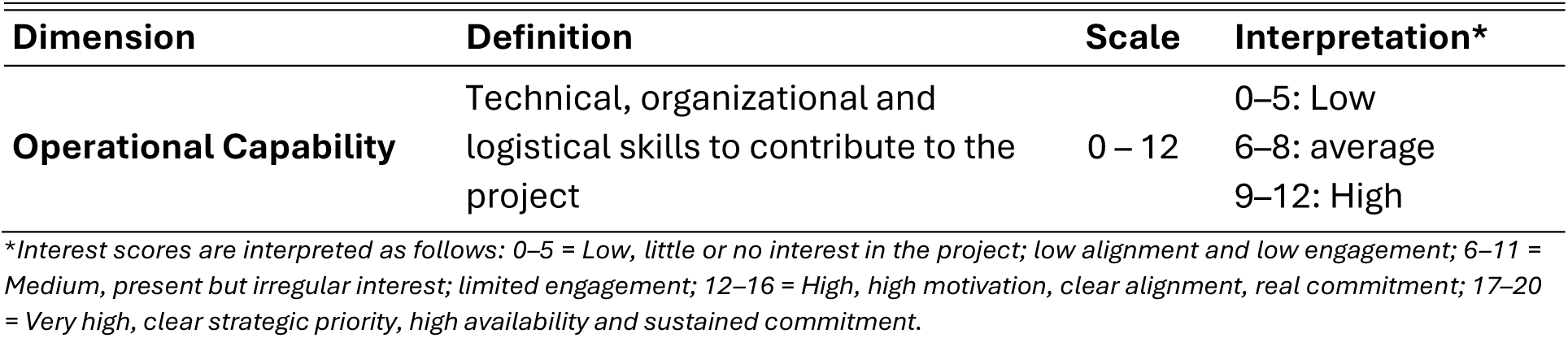
Categorization of Stakeholders according to their average score of interest-power-operational capacity.

**Table 2b:** Categorization of stakeholders in Mali and Burkina Faso according to their power and influence in the prevention and control of genital chlamydia among young people aged 15-24 years.

| Score Total | Level | Definition (Power/Influence) |
| --- | --- | --- |
| **0–3** | **Low** | Little influence on policies, resources or networks. |
| **4–7** | **Medium** | Moderate influence, capacity to act but limited. |
| **8–10** | **High** | Significant influence on resources, guidance and networks. |
| **11–12** | **Very high** | Key player, major strategic influence, strong decision-making power. |

**Table 2c:** Categorization of stakeholders in Mali and Burkina Faso according to their operational capacity in the prevention and control of genital chlamydia in young people aged 15-24 years.

| Score Total | Level | Definition (Operational Capability) |
| --- | --- | --- |
| **0–3** | **Low** | Low expertise, limited coverage, and poorly developed M&E system. |
| **4–7** | **Medium** | Existing but partial or inconsistent capacities. |
| **8–10** | **High** | Good expertise, extensive coverage, functional M&E. |
| **11–12** | **Very high** | Excellent capabilities, strong expertise, national coverage, robust M&E. |

#### 2.5.2. Data Analysis

Consolidated scores were descriptively analyzed. For each stakeholder and dimension studied, the means and standard deviations of the scores assigned by five (5) independent evaluators were calculated (**Table 3a**).

**Table 3a:** Average stakeholder scores (interest, influence, and capacity) in genital chlamydia prevention among youths aged 15–24 years old in Mali and Burkina Faso.

| # | Stakeholder | Interest ($\mu \pm \sigma$) | Power ($\mu \pm \sigma$) | Implementation Capacity ($\mu \pm \sigma$) |
| --- | --- | --- | --- | --- |
| 1 | RJASR/PF (ML) | 19.0 ± 2.0 | 9.0 ± 2.4 | 11.0 ± 1.6 |
| 2 | SE/HCNLS (ML) | 17.2 ± 1.9 | 10.8 ± 2.3 | 10.8 ± 1.3 |
| 3 | PSSLS-IST/HV (BF) | 17.0 ± 2.3 | 10.6 ± 1.7 | 10.4 ± 1.1 |
| 4 | SP/CNLS-IST (BF) | 17.0 ± 1.6 | 11.5 ± 1.0 | 10.8 ± 1.2 |
| 5 | CSLS-TBH (ML) | 17.0 ± 1.6 | 11.5 ± 1.0 | 10.8 ± 1.2 |
| 6 | REGIPIV-BF (BF) | 16.0 ± 1.5 | 07.8 ± 0.8 | 10.6 ± 1.7 |
| 7 | BURCASO (BF) | 16.0 ± 3.0 | 07.8 ± 1.2 | 09.5 ± 1.9 |
| 8 | ENDA Mali (ML) | 15.6 ± 1.8 | 06.4 ± 2.7 | 08.6 ± 2.8 |
| 9 | RNPEM (ML) | 13.0 ± 2.5 | 05.0 ± 2.6 | 08.0 ± 1.6 |
| 10 | NGO Muso (ML) | 13.0 ± 2.5 | 05.0 ± 1.5 | 08.0 ± 1.6 |

\*ML= Mali BF= Burkina Faso NGO ENDA Mali: the Malian national branch of the Enda Third World network BURCASO: Burkinabe Council of Community Development Organizations CSLS-TBH: Sectoral Unit for the Fight against HIV/AIDS, Tuberculosis and Viral Hepatitis REGIPIV-BF: Network for a Greater Involvement of People Infected and Affected by HIV/HIV-AIDS RJASR/PF: Network of Young Ambassadors for Reproductive Health/Family Planning RNPEM: National Network of Peer Educators of Mali PSSLS-IST/HV: Health Sector Programme for the Fight against AIDS and Sexually Transmitted Infections and Viral Hepatitis SP/CNLS-IST: Permanent Secretariat of the National Council for the Fight against AIDS and STIs SE/HCNLS: Executive Secretariat of the High National Council for the Fight against AIDS

**Table 3b:** Coefficient of variation (CV%) of scores assigned to stakeholders.

| # | Stakeholders | Coefficient of variation of average CV scores (%) |  |  |
| --- | --- | --- | --- | --- |
|  |  | Interest | Power | Capacity |
| 1 | RNPEM (ML) | 19.2 % | 52.0 % | 20.0 % |
| 2 | NGO Muso (ML) | 19.2 % | 30.0 % | 20.0 % |
| 3 | BURCASO (BF) | 18.8 % | 15.4 % | 20.0 % |
| 4 | PSSLS-IST/HV (BF) | 13.5 % | 16.0 % | 10.6 % |
| 5 | NGO ENDA Mali (ML) | 11.5 % | 42.2 % | 32.6 % |
| 6 | SE/HCNLS (ML) | 11.0 % | 21.3 % | 12.0 % |
| 7 | RJASR/PF (ML) | 10.5 % | 26.7 % | 14.5 % |
| 8 | REGIPIV-BF (BF) | 09.4 % | 10.3 % | 16.0 % |
| 9 | CSLS-TBH (ML) | 09.4 % | 08.7 % | 11.1 % |
| 10 | SP/CNLS-IST (BF) | 09.4 % | 08.7 % | 11.1 % |

\*ML= Mali BF= Burkina Faso NGO ENDA Mali: the Malian national branch of the Enda Third World network BURCASO: Burkinabe Council of Community Development Organizations CSLS-TBH: Sectoral Unit for the Fight against HIV/AIDS, Tuberculosis and Viral Hepatitis REGIPIV-BF: Network for a Greater Involvement of People Infected and Affected by HIV/HIV-AIDS RJASR/PF: Network of Young Ambassadors for Reproductive Health/Family Planning RNPEM: National Network of Peer Educators of Mali PSSLS-IST/HV: Health Sector Programme for the Fight against AIDS and Sexually Transmitted Infections and Viral Hepatitis SP/CNLS-IST: Permanent Secretariat of the National Council for the Fight against AIDS and STIs SE/HCNLS: Executive Secretariat of the High National Council for the Fight against AIDS

We calculated the coefficient of variation (CV) to assess the agreement between scores from stakeholder assessments using the following formula: CV=(μ/σ) X 100 with a CV < 10% (strong agreement), between 10–20% (moderate agreement) and > 20% (low agreement) (**Table 3b**).

Data entry and processing were carried out using Microsoft Excel 2019 software. Based on the consolidated scores, stakeholders were grouped into three (3) categories: institutional actors, technical actors and operational actors (NGOs). The results were graphed in the form of Mendelow matrices crossing the interest, power and operational capacity of the stakeholders (**Figure 1**).

**Figure 1:**
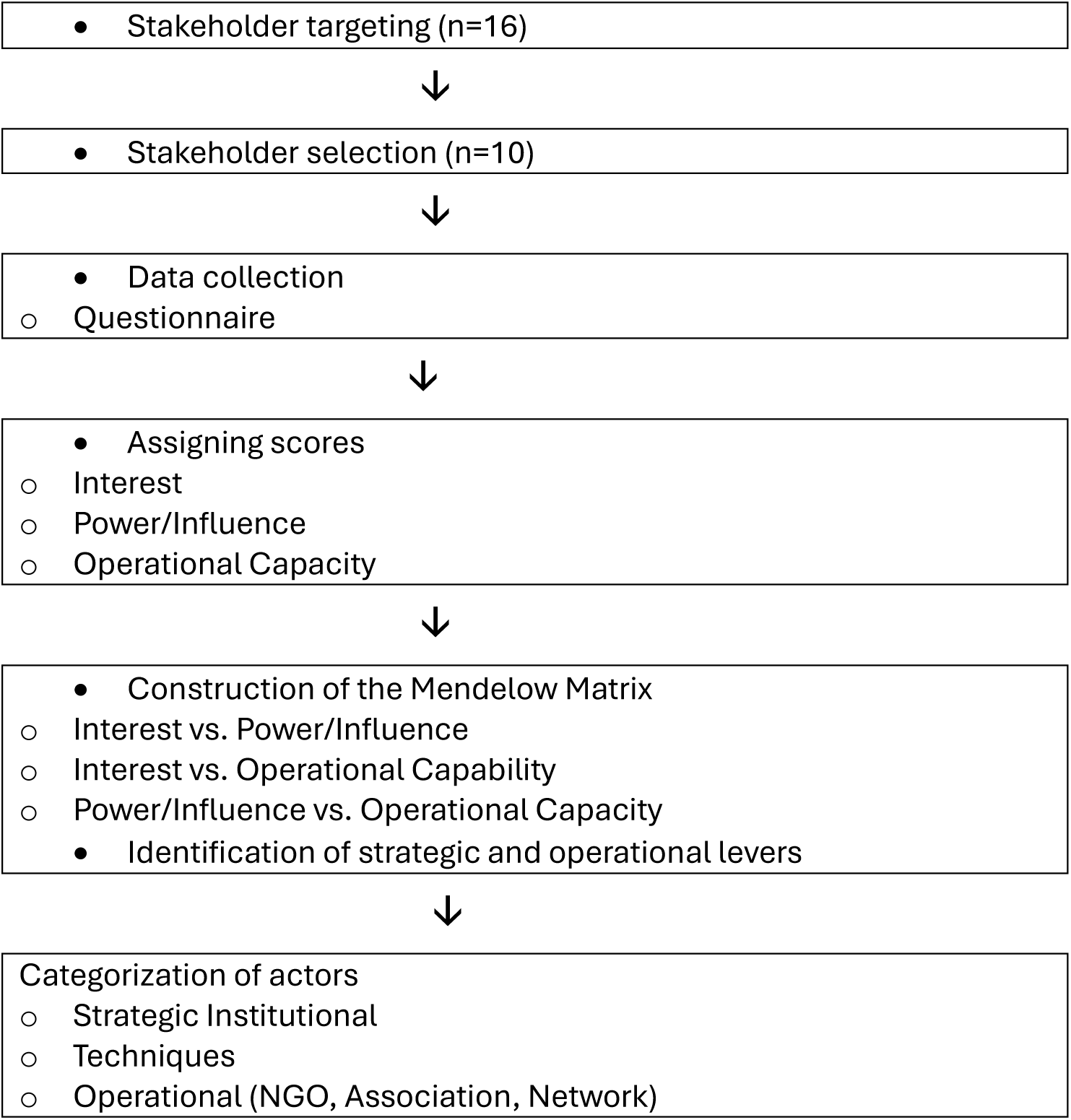
Process for selecting, evaluating, and classifying stakeholders involved in the prevention and control of genital chlamydia among young people aged 15–24 years in Mali and Burkina Faso

### 2.6. Ethical considerations

The study protocol, questionnaire, and informed consent forms were approved by the Ethics Committee of the Mali Hospital in Bamako (Reference No. 2025-001) and by the Ethics Committee of the African Institute of Public Health (IASP) in Ouagadougou (Reference No. 2025-06-230). Informed consent was obtained from participants prior to participation. The data collected did not contain any nominative or sensitive information. Personal identifiers were removed, and only the principal investigator had access to the raw data. The confidentiality and anonymity of participants and evaluators were guaranteed throughout the process. Symbolic monetary compensation was granted to the participants.

## 3. RESULTS

A total of ten (10) responses were received, including six (6) in Mali and four (4) in Burkina Faso.

**Table 4** highlights a sample of experienced professionals in strategic positions with strong thematic expertise, ensuring the relevance and credibility of the information collected.

Table 3a presents the average scores (μ ± σ) assigned to the ten stakeholders involved in the prevention and control of STIs, including genital chlamydia in young people aged 15–24 years, according to the three dimensions assessed: interest, power/influence, and operational capacity.

### 3.1. Average scores of the three dimensions assessed

#### 3.1.1. Level of Interest

Interest scores range from 13.0/20 to 19.0/20, indicating an overall high level of engagement among stakeholders. The highest score was observed in RJASR/PF (19.0 ± 2.0). National coordination structures such as the SE/HCNLS (17.2 ± 1.9), the SP/CNLS-IST (17.0 ± 1.6) and the CSLS-TBH (17.0 ± 1.6) also showed similarly high levels of interest. Community-based NGOs, including ENDA Mali (15.6 ± 1.8), BURCASO (16.0 ± 3.0) and REGIPIV-BF (16.0 ± 1.5), showed a lower level of interest. The lowest scores are observed for the RNPEM (13.0) and the NGO Muso (13.0), corresponding to a moderate level of interest (**Table 3a**).

#### 3.1.2. Power / Influence

The average power/influence scores showed greater dispersion, ranging from 5.0/12 to 11.5/12. The highest levels are recorded in SP/CNLS-IST and CSLS-TBH (11.5 ± 1.0), followed by SE/HCNLS (10.8 ± 2.3). NGOs and community networks have more moderate levels of influence: the NGO ENDA Mali (6.4 ± 2.7), BURCASO (7.8 ± 1.2) and REGIPIV-BF (7.8 ± 0.8). The lowest scores are observed for the RNPEM and the NGO Muso (5.0).

#### 3.1.3. Operational Capability

The average operational capability scores were relatively homogeneous, ranging from 8.0/12 to 11.0/12. RJASR/PF (Mali) obtained the highest score (11.0 ± 1.6). National institutions (SP/CNLS-IST, CSLS-TBH, SE/HCNLS and PSSLS-IST/HV) and community-based organizations such as REGIPIV-BF (10.6 ± 1.7) and PSSLS-IST/HV (10.4 ± 1.1) also showed high levels of capacity (≈10.8/12). The lowest scores were for the RNPEM and the NGO Muso (8.0).

### 3.2. Coefficients of variation between valuations

Table **3b** presents the coefficients of variation (CV%) of the evaluators’ scores to the ten stakeholders according to the three dimensions analyzed: interest, power/influence, and operational capacity.

#### 3.2.1. Interest Level

CVs for interest range from 9.4% to 19.2%, indicating an overall low to moderate dispersion of scores. The SP/CNLS-IST, CSLS-TBH and REGIPIV-BF showed the lowest levels of variability (≈9.4%), reflecting strong inter-rater agreement. The highest levels of variability were observed for RNPEM and the NGO Muso (19.2%) (**Table 3b).**

#### 3.2.2. Power / Influence

The power/influence dimension had the highest dispersion of evaluations. The PUCs range from 8.7% to 52.0%. The SP/CNLS-IST and the CSLS-TBH had the lowest CVs (8.7%). The RNPEM (52.0%), the NGO ENDA Mali (42.2%) and the NGO Muso (30.0%) showed high levels of variability (**Table 3b).**

#### 3.2.3. Operational Capability

The CVs for operational capability range from 10.6% to 32.6%. SP/CNLS-STI, CSLS-TBH, PSSLS-IST/HV showed low to moderate dispersion (≈10–11%). The NGO ENDA Mali (32.6%) showed more marked variability (**Table 3b).**

### 3.3. Cross-Dimensional Analysis (Mendelow Matrices)

#### 3.3.1. Power-Interest Analysis (**Figure 2a**)

The Power-Interest matrix identified four (4) profiles:

◦ High power – strong interest: SP/CNLS-IST, CSLS-TBH, SE/HCNLS and PSSLS-IST/HV.
◦ High interest – moderate/weak power: RJASR/PF, REGIPIV-BF, BURCASO and the NGO ENDA Mali.
◦ Low power – low interest: the RNPEM and the NGO Muso.
◦ High Power – Low Interest: No Stakeholder.

**Figure 2a:**
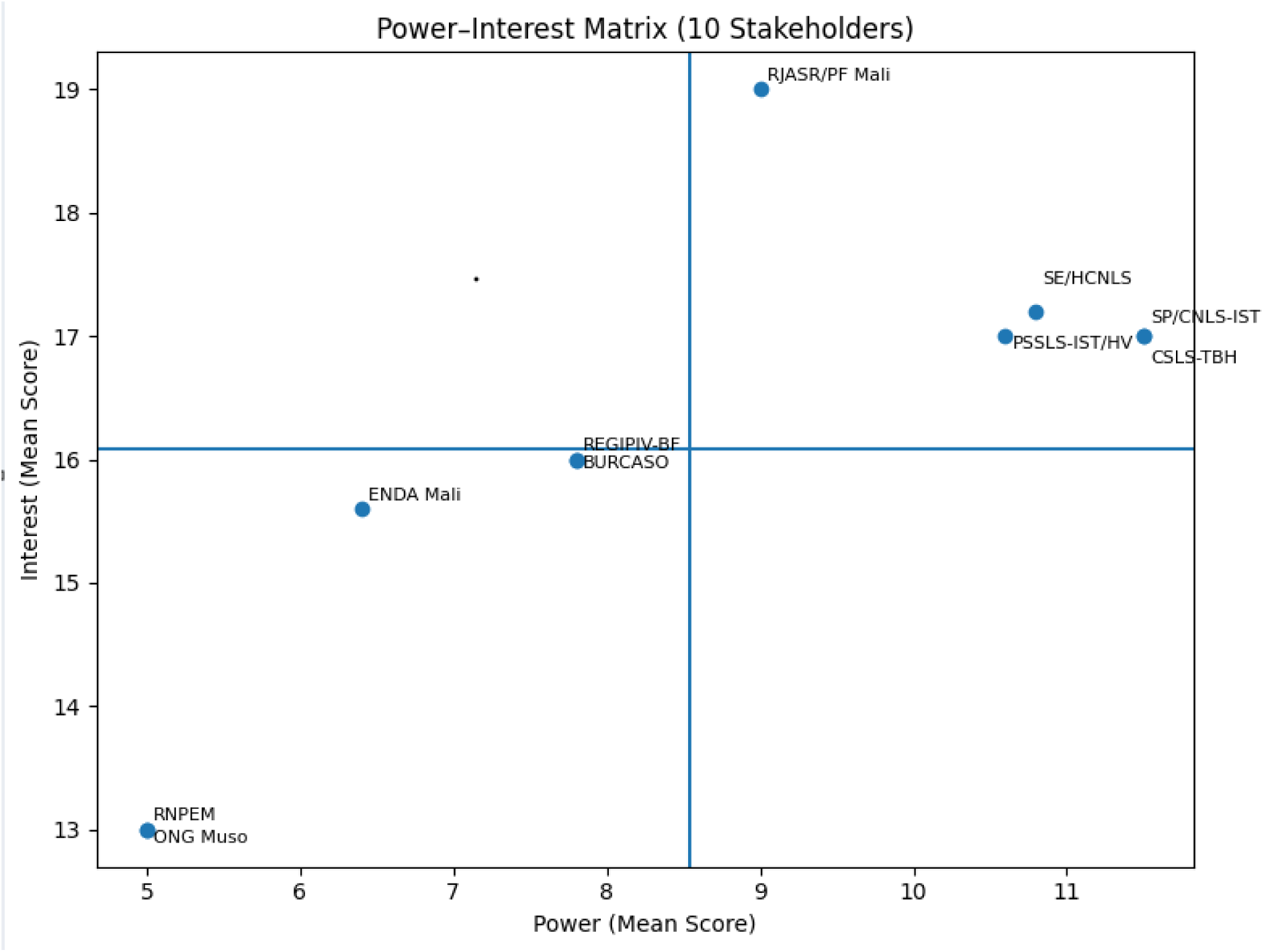
Power-interest analysis of stakeholders in the prevention and control of genital chlamydia in young people aged 15-24 in Mali and Burkina Faso

**Figure 2b:**
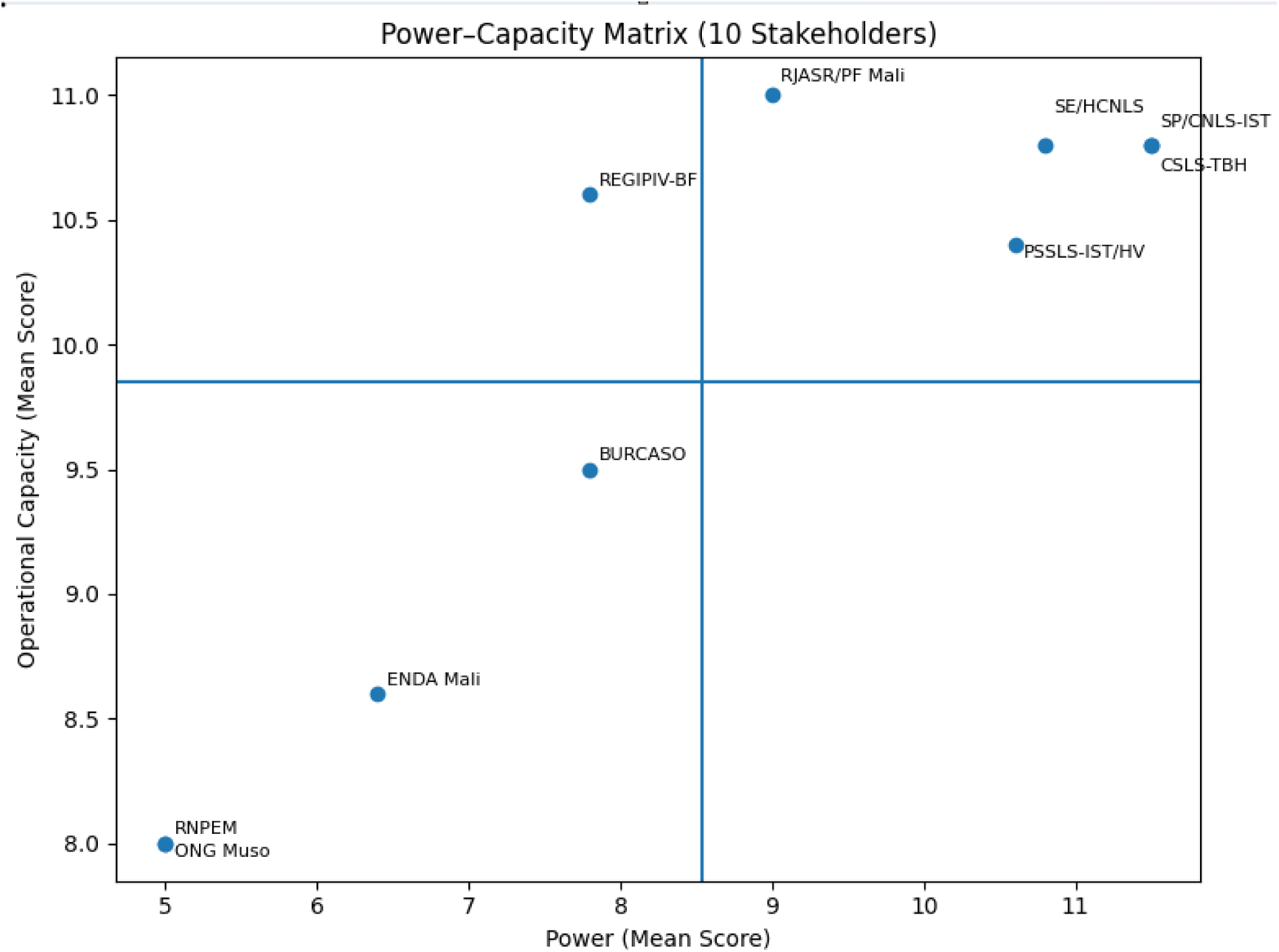
Power Analysis – Operational Capacity of Stakeholders in the Prevention and Control of Genital Chlamydia in Young People Aged 15-24 in Mali and Burkina Faso

**Figure 2c:**
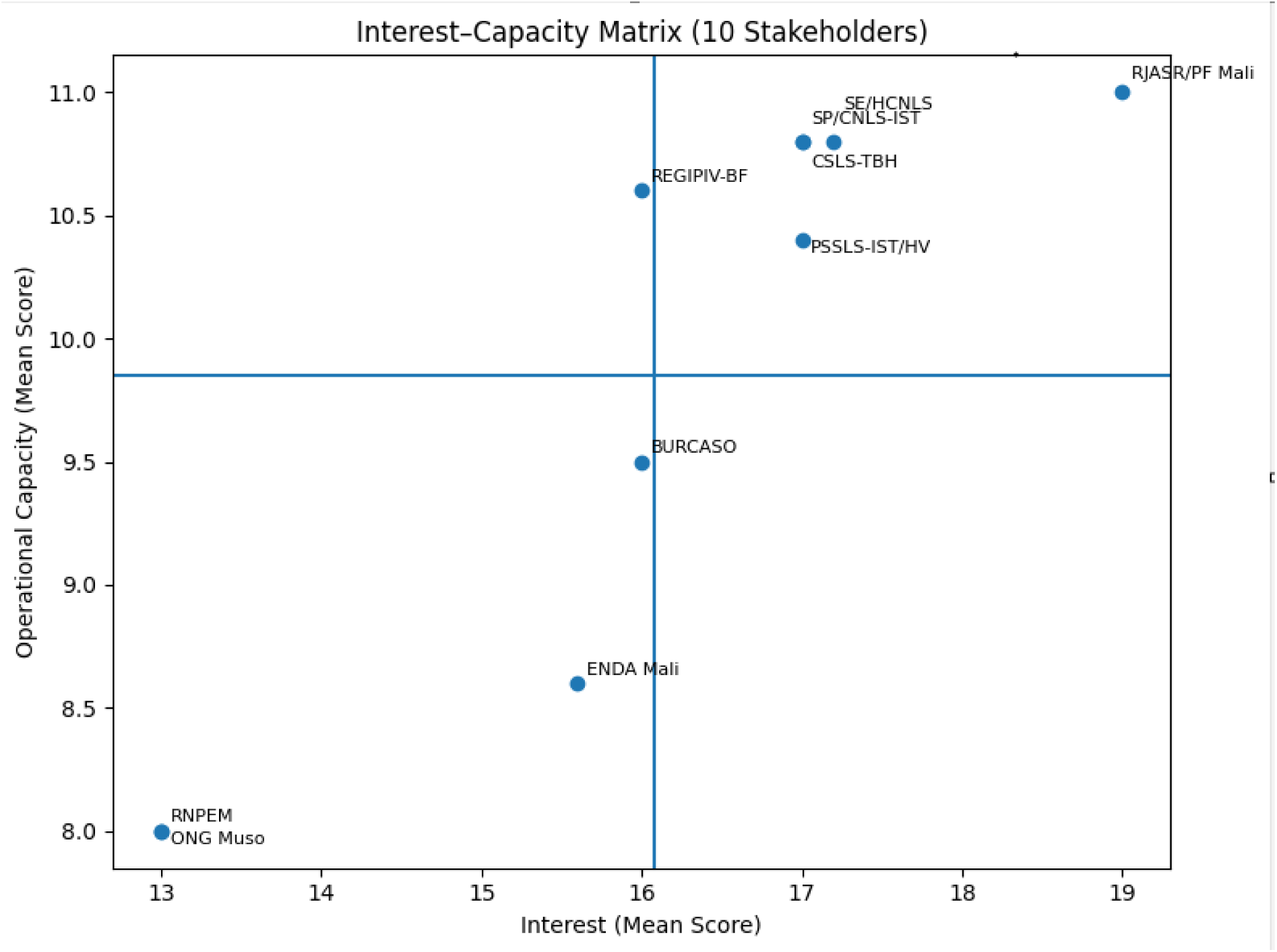
Interest-Operational Capacity Analysis of Stakeholders in the Prevention and Control of Genital Chlamydia in Young People Aged 15-24 in Mali and Burkina Faso

#### 3.3.2. Analysis Authority – Operational Capability (Figure 2b)

The Power-Operational Capability matrix highlighted four (4) profiles:

◦ High power – strong capacity: National institutions (SP/CNLS-IST, CSLS-TBH, SE/HCNLS, PSSLS-IST/HV).
◦ Low power – low capacity: Community-based organizations and networks (RJASR/PF, REGIPIV-BF, BURCASO, NGO ENDA Mali).
◦ Low power – moderate to low capacity: RNPEM and the NGO Muso.
◦ High Power – Low Capacity: No Stakeholder.

#### 3.3.3. Interest Analysis – Operational Capability (Figure 2c)

The Interest–Ability Matrix also revealed four (4) profiles:

◦ High Interest - High Capacity: SP/CNLS-IST, CSLS-TBH, SE/HCNLS, PSSLS-IST/HV, and RJASR/PF
◦ High interest - moderate capacity: REGIPIV-BF, BURCASO and the NGO ENDA Mali
◦ Low interest – low capacity: RNPEM and the NGO Muso
◦ Low Interest – High Capacity: No Stakeholder.

### 3.4. Strategy for engaging institutional and community partners in the project

Table 4 presents a structured mapping of key stakeholders to be involved in the sustainability and scaling of the Pfizer FAPH chlamydia project in Mali and Burkina Faso. At the institutional level, the national structures for the fight against HIV/AIDS and STIs (SE/HCNLS in Mali, SP/CNLS-IST in Burkina Faso), as well as the specialized sectoral units and programs (PSSLS-IST/HV in Burkina Faso and the CSLS-TBH in Mali) will mainly be engaged through formal exchanges, often online, by email, telephone or during seminars. Interactions will generally be annual to quarterly and focus on strategic dialogue, technical coordination, data transmission, monitoring of indicators, as well as requesting funding and sharing results and reports. CSOs and community networks (NGOs, national networks and associations) will favour more flexible formats, combining face-to-face meetings (often by appointment) and online exchanges. Means of communication will include email, telephone and videoconferencing platforms. The frequency of exchanges varies from monthly to several times a year, depending on operational needs. The objectives will focus on the coordination of field activities, monitoring of community interventions, community mobilization, business planning, M&E information sharing, as well as advocacy.

**Table 4:**
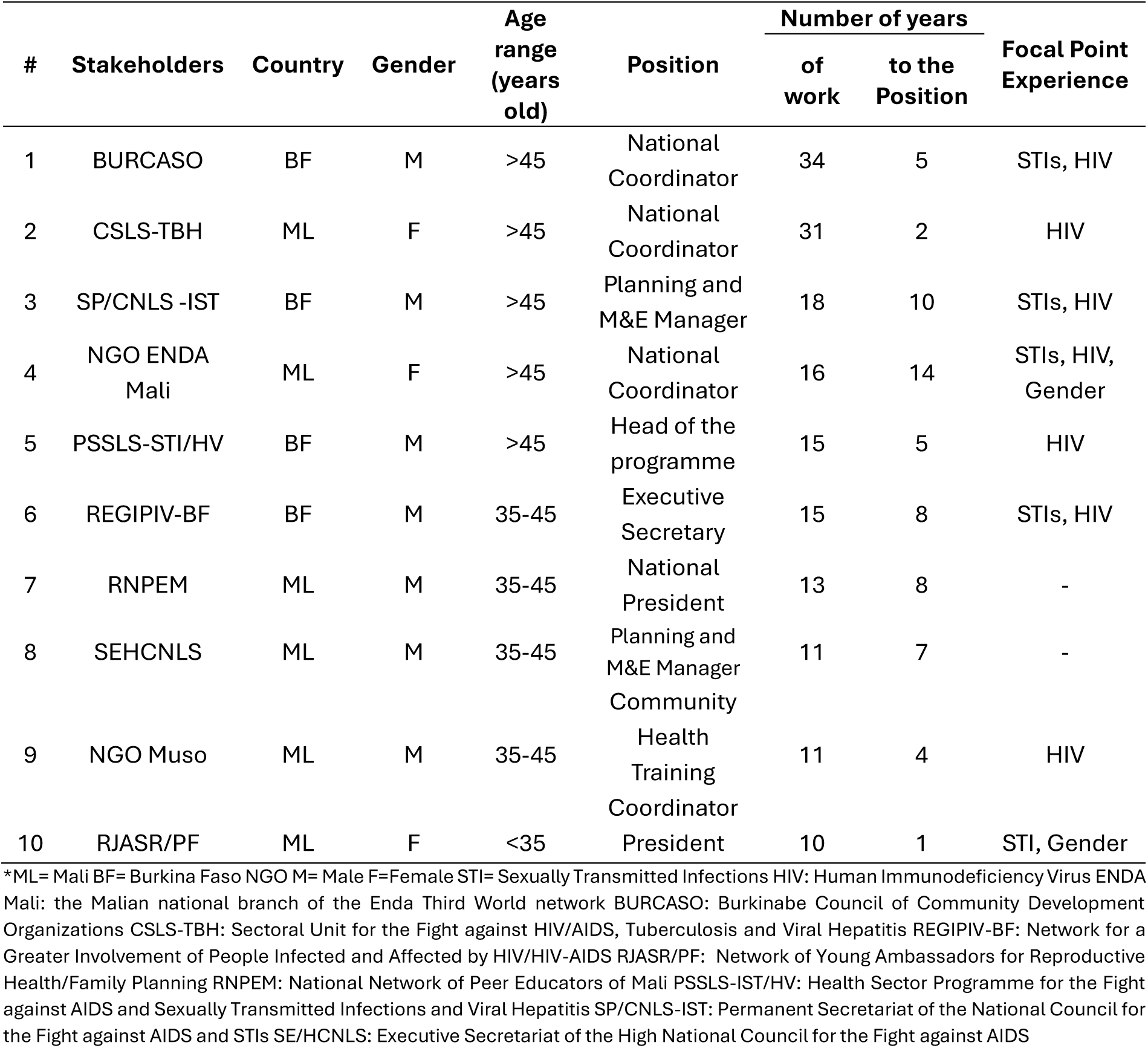
Description of Respondents by Stakeholder in Mali and Burkina Faso.

**Table 5:**
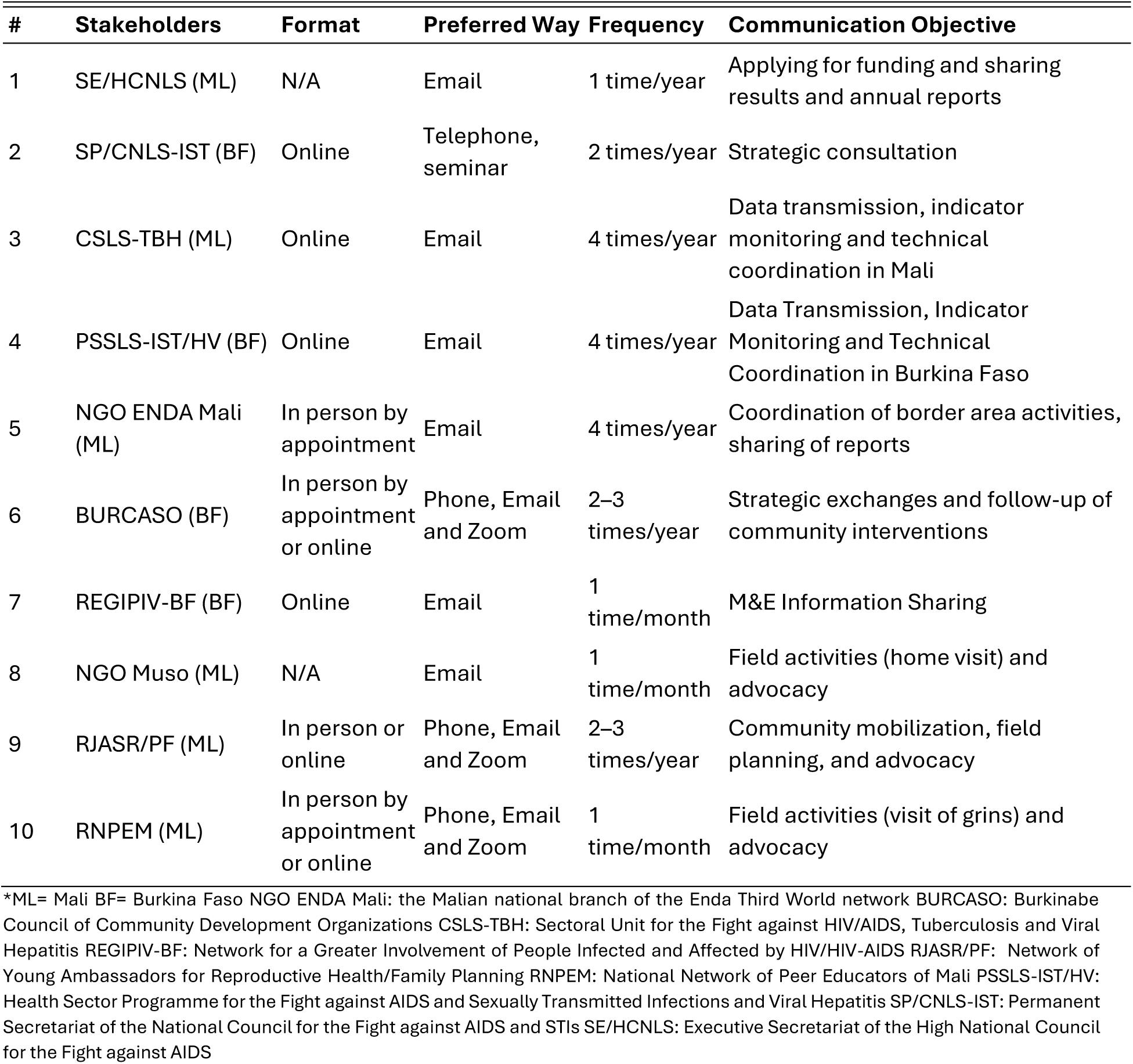
Information for the development of the Institutional and Community Partner Engagement Strategy for the project in Mali and Burkina Faso.

| # | Stakeholders | Format | Preferred Way | Frequency | Communication Objective |
| --- | --- | --- | --- | --- | --- |
| 1 | SE/HCNLS (ML) | N/A | Email | 1 time/year | Applying for funding and sharing results and annual reports |
| 2 | SP/CNLS-IST (BF) | Online | Telephone, seminar | 2 times/year | Strategic consultation |
| 3 | CSLS-TBH (ML) | Online | Email | 4 times/year | Data transmission, indicator monitoring and technical coordination in Mali |
| 4 | PSSLS-IST/HV (BF) | Online | Email | 4 times/year | Data Transmission, Indicator Monitoring and Technical Coordination in Burkina Faso |
| 5 | NGO ENDA Mali (ML) | In person by appointment | Email | 4 times/year | Coordination of border area activities, sharing of reports |
| 6 | BURCASO (BF) | In person by appointment or online | Phone, Email and Zoom | 2–3 times/year | Strategic exchanges and follow-up of community interventions |
| 7 | REGIPIV-BF (BF) | Online | Email | 1 time/month | M&E Information Sharing |
| 8 | NGO Muso (ML) | N/A | Email | 1 time/month | Field activities (home visit) and advocacy |
| 9 | RJASR/PF (ML) | In person or online | Phone, Email and Zoom | 2–3 times/year | Community mobilization, field planning, and advocacy |
| 10 | RNPEM (ML) | In person by appointment or online | Phone, Email and Zoom | 1 time/month | Field activities (visit of grins) and advocacy |

\*ML= Mali BF= Burkina Faso NGO ENDA Mali: the Malian national branch of the Enda Third World network BURCASO: Burkinabe Council of Community Development Organizations CSLS-TBH: Sectoral Unit for the Fight against HIV/AIDS, Tuberculosis and Viral Hepatitis REGIPIV-BF: Network for a Greater Involvement of People Infected and Affected by HIV/HIV-AIDS RJASR/PF: Network of Young Ambassadors for Reproductive Health/Family Planning RNPEM: National Network of Peer Educators of Mali PSSLS-IST/HV: Health Sector Programme for the Fight against AIDS and Sexually Transmitted Infections and Viral Hepatitis SP/CNLS-IST: Permanent Secretariat of the National Council for the Fight against AIDS and STIs SE/HCNLS: Executive Secretariat of the High National Council for the Fight against AIDS

## 4. DISCUSSION

The analysis of the stakeholders of the Pfizer Chlamydia Project FAPH highlighted the diversity and complementarity of the actors involved in the prevention and control of genital chlamydia in young people aged 15 to 24 in Mali and Burkina Faso.

Respondents on behalf of stakeholders were over 45 years of age (5), followed by those aged 35 to 45 (4) and one person under 35 years of age, reflecting a high level of professional experience. The respondents mainly held positions of high responsibility (national coordination, planning and monitoring and evaluation, medical care and associative leadership), thus targeting key decision-making actors. Professional seniority ranges from 10 to 34 years, with experience in the position ranging from 1 to 14 years. Most of them had focal point expertise in STIs and HIV, sometimes gender-mainstreaming, attesting to their strong technical and strategic competence (**Table 4**).

### 4.1. Stakeholder Analysis in Three Dimensions (Interest, Power/Influence, Capacity)

This approach to stakeholder analysis in the three (3) dimensions (interest, power/influence, and operational capacity) is a strategic tool to identify operational levers and potential constraints, particularly in terms of coordination, resource mobilization and M&E. It can guide future planning of similar programs in other West African contexts, ensuring the sustainability and scale-up of interventions targeting STIs in youth (Rowley et al., 2019; UNFPA, 2021).

National institutions SP/CNLS-IST in Burkina Faso and SE/HCNLS in Mali have distinguished themselves by high interest and power, reflecting their central role in the coordination and strategic planning of sexual and reproductive health programs (Rowley et al., 2019; WHO, 2021). The technical structures CSLS-TBH in Mali and PSSLS-IST/HV in Burkina Faso have shown a strong operational capacity, essential for technical support, surveillance and integration of screening and monitoring activities (UNFPA, 2021).

The major strategic players were the SP/CNLS-IST, CSLS-TBH and SE/HCNLS combining high interest, strong power and strong operational capacity. The key operational actors (civil society and networks) were RJASR/PF, REGIPIV-BF and BURCASO, characterized by strong interest and capacity, but more moderate power. The actors with low influence were the RNPEM and the NGO Muso, which had lower scores in all three dimensions **(Table 3a).**

NGOs and community networks, such as the NGO ENDA Mali, BURCASO and REGIPIV-BF, have a high level of interest but moderate power, reflecting their engagement on the ground and their key role in community mobilization, but their influence on strategic decisions remains limited. These findings are consistent with previous observations on the importance of CSOs in accessing young and STI-vulnerable populations in sub-Saharan African countries (Hu et al., 2025; Michalow et al., 2025).

### 4.2. Coefficient of variation and concordance of stakeholder assessments

An indirect analysis of inter-rater agreement, based on the coefficient of variation of the scores assigned, indicates an overall satisfactory homogeneity of the evaluations, particularly for national institutions whose coefficients of variation are less than 10% for the power dimension. On the other hand, a greater dispersion is observed for some community organizations, particularly for the power/influence dimension, suggesting a more heterogeneous perception of their strategic position. Overall, these results reflect moderate to strong inter-rater agreement, reinforcing the robustness of the consolidated scores.

The inter-evaluator concordance appears satisfactory for the interest and operational capacity dimensions, particularly within national institutions. The power/influence dimension is characterized by greater variability, especially for community-based organizations, which probably reflects the more subjective nature of the evaluation of strategic influence. Despite this finding, the evaluation of the dimensions of interest, power and operational capacity shows that the success and sustainability of the project depend on inclusive governance, combining institutional leadership, technical expertise, and community engagement. Indeed, the active involvement of actors at all levels not only ensures the effectiveness of interventions, but also strengthens the resilience of local sexual and reproductive health systems (Sameni et al., 2025; WHO, 2021).

### 4.3. Matrix Analysis

#### 4.3.1. Power – Interest

The power–interest analysis showed an overall favorable configuration for the governance of the prevention and control of genital chlamydia among young people aged 15-24 years in Mali and Burkina Faso (**Figure 2a**).

The national coordinating institutions (the SP/CNLS-IST, CSLS-TBH, SE/HCNLS, PSSLS-IST/HV) which have accumulated strong power and strong interest. This reflects a strategic alignment between decision-making authority and thematic commitment, which is an essential condition for promoting clear political orientations, mobilising human and financial resources and ensuring the follow-up of interventions. Their positioning will strengthen the institutional coherence and legitimacy of the chlamydia project activities in case of sustainability and/or scale-up.

NGOs and community networks (RJASR/PF, REGIPIV-BF, BURCASO, NGO ENDA Mali) have high interest and limited power. This situation indicates that their operational dynamics are largely based on actors committed to the field, particularly with young people, but dependent on the decisions and funding of national authorities. This creates a need for capacity building, advocacy and more formal integration into decision-making mechanisms in order to maximize their impact.

The RNPEM and the NGO Muso, classified as low power – low interest, occupy a marginal position in strategic governance. This may reflect either a current low level of involvement or a perceived mismatch between their mandate and the issue. Raising awareness or clarifying roles could increase their contribution.

No actor was of high power – low interest. This is particularly positive because it suggests an institutional convergence around the importance of the fight against genital chlamydia in young people, reducing the risk of political or institutional blockage of the project.

#### 4.3.2. Authority – Operational Capability

The Power–Operational Capacity analysis highlights a globally balanced and functional institutional architecture in the fight against genital chlamydia among young people aged 15-24 in Mali and Burkina Faso (**Figure 2b**).

The national structures (the SP/CNLS-IST, CSLS-TBH, SE/HCNLS, PSSLS-IST/HV) show strong power and operational capacity. This reflects a coherence between decision-making authority and technical skills, reducing the risk of a mismatch between strategic planning and implementation across CSOs or beneficiaries. Their central position will promote effective governance and structured coordination of the project.

Community-based organizations (RJASR/PF, REGIPIV-BF, BURCASO, NGO ENDA Mali) have a strong capacity for implementation, especially with young people, but limited institutional power. This configuration suggests a dependence on the orientations and resources of the national authorities. The strategic challenge is therefore to strengthen their institutional recognition and their participation in decision-making processes, in order to optimize the impact of interventions.

The RNPEM and the NGO Muso, classified as low power – low capacity, occupy a marginal position. This indicates an under-explored potential or a weak integration into the system. Targeted capacity-building and integration actions could improve overall synergy.

No player is in a situation of high power – low capacity, which is a very positive signal. This means that policymakers also have the technical means to operationalize policies, limiting the risks of institutional inefficiency.

#### 4.3.3. Interest – Operational Capability

The Interest–Operational Capacity analysis reveals a good coherence between the commitment of actors and their ability to act in the fight against genital chlamydia among young people aged 15-24 years (**Figure 2c**).

The national institutions (SP/CNLS-IST, CSLS-TBH, SE/HCNLS, PSSLS-IST/HV) as well as RJASR/PF combine strong interest and capacity. This configuration is strategic: it means that the most motivated actors also have the technical and organizational resources necessary to effectively implement interventions. They thus constitute the operational pillars of the system.

REGIPIV-BF, BURCASO and the NGO ENDA Mali show a high level of interest but a moderate capacity. Their commitment is a major asset, but technical, logistical or financial reinforcement could improve their performance and broaden their impact with young people.

The RNPEM and the NGO Muso, positioned in low interest – low capacity, play a limited role. This may reflect poor strategic alignment or a lack of resources. Awareness-raising and strengthening actions could promote their integration.

No player is in low interest – high capacity. This is positive, because there are no significant technical resources that are insufficiently committed to the problem.

Taken together, the three (3) matrices revealed a coherent system where the decision-makers (the SP/CNLS-IST, CSLS-TBH, SE/HCNLS, PSSLS-IST/HV) are committed and technically competent, while the community actors (the REGIPIV-BF, the BURCASO, and the NGO ENDA Mali) are motivated and operationally active, but less influential at the institutional level. The most committed actors generally have a good capacity for action, which is a major asset in the fight against STIs among young people. The main strategic challenge lies in strengthening the capacities of the less well-endowed CSOs (RJASR/PF, RNPEM and the NGO Muso) and in consolidating the bridges between decision-making power and field expertise, in order to ensure an integrated, participatory and sustainable coordination of the project’s interventions.

In sexual and reproductive health programmes in sub-Saharan Africa, there has been a concentration of decision-making power at the level of national institutions, combined with a strong operational capacity of community-based organizations and civil society networks (UNFPA, 2021; CSLS-TBH, 2020; CNLS-IST, 2020; Rowley et al., 2019). This asymmetrical distribution of power is frequently described in STI and HIV programmes, where state structures provide strategic coordination and resource mobilisation, while NGOs play a central role in outreach implementation and access to vulnerable populations.

The definition of a clear strategy for engaging stakeholders is an operational tool for planning and harmonizing communication and collaboration with institutional and community actors, specifying for each the format of interaction, the preferred means of communication, the frequency of exchanges and the objectives sought.

### 4.4. Limitations of the study

This study has limitations. Data collection was based on self-administered questionnaires, which can introduce biases related to the subjectivity of responses or overestimation of participants’ interest and engagement. The assessment of power and operational capacity was based on scores assigned by evaluators, which, despite their independence, remains partially subjective. Finally, the study did not integrate the perspective of the final beneficiaries (young people aged 15–24), which limits a full understanding of the implementation dynamics on the ground.

## 5. CONCLUSION

This study made it possible to clearly identify the key actors involved in the prevention and control of genital chlamydia in young people aged 15 to 24, as well as the operational relays and the main levers for action. The power-interest matrix highlights a hierarchical but complementary architecture of actors, characterized by a strong institutional core providing leadership and strategic coordination, and a structured community network, which is essential for the implementation of outreach interventions to young people.

The analysis of the positioning and level of stakeholder engagement revealed a configuration that is generally favorable to the implementation, sustainability and scale-up of the Pfizer Chlamydia Project led by the Faculty of Pharmacy (FAPH). Actors of high interest generally have adequate operational capacities, while close coordination between public institutions and community-based organizations appears to be a determining factor in strengthening the effectiveness of project interventions or activities in Mali and Burkina Faso.

## 6. PERSPECTIVES

Five (5) strategic orientations were considered to enable the sustainability and scale-up of the Pfizer Chlamydia Project led by the Faculty of Pharmacy (FAPH).

◦ The institutional leadership of the project will be strengthened through the mobilization of high-power and high-capacity actors, including the SE/HCNLS in Mali and the SP/CNLS-IST in Burkina Faso, in governance bodies, multisectoral coordination and strategic monitoring, to ensure sustainable alignment with national sexual and reproductive health policies.
◦ Community-based organizations with a strong interest and operational capacity, such as NGO ENDA Mali and REGIPIV-BF, will be positioned as priority operational partners for the implementation of outreach interventions to young people aged 15 to 24.
◦ Actors with a strong interest but limited power or capacity, such as RJASR/PF, will benefit from targeted advocacy and capacity building support in the management of adolescent and youth reproductive health (SRAJ) projects.
◦ Existing specific operational skills, in particular home visits (VAD) and *grin* visits, will be valued with actors with low power and low influence, such as the RNPEM and NGO Muso, in order to optimize the reach of the target populations.
◦ In the short and medium term, all stakeholders will be in the project’s steering committee but the commitment will cover three (3) to four (4) key stakeholders maximum. An annual evaluation of the power-interest, power-to-capacity, and interest-capacity matrices will be carried out in Mali and Burkina Faso in order to update the prioritization of actors to be engaged and to adjust the strategic objectives of the project.

## Data Availability

All data produced in the present work are contained in the manuscript.

## 8. ANNEXES

### 8.1. 1-page Summary sheet template

Each stakeholder’s one-page summary sheet included information on the following evaluation criteria with operational definitions:

#### 1. Interest

◦ Strategic alignment: The degree to which the stakeholder is interested in the prevention and management of STIs and genital chlamydia in young people aged 15-24 years.
◦ Operational commitment: The willingness of the stakeholder to provide human and technical resources for the said project in the future.
◦ Sustainability: The willingness of the stakeholder to integrate the results of the project into national policies and programmes (coordination level) or into its activities or interventions in the field (operational level).
◦ Action plan: The existence, clarity and relevance of the actions planned by the stakeholder to contribute to the project in the short, medium and long term
◦ Availability for the Pfizer FAPH chlamydia project: the degree of real and immediate mobilization of the stakeholder to support the project, including the institutional priority given to the project, the availability of available staff and skills, the ability to devote time to activities, as well as the possibility of quickly allocating financial resources (co-financing for example).

#### 2. Power/Influence

◦ Decision-making power: the ability to influence national STI policies, budgets and orientations among young people aged 15-24.
◦ Resource control: Access to the national budget, TFP financing and infrastructure for the said project.
◦ Networking and partnerships: The ability to mobilize other TFPs or network with other actors in the fight against STIs (patient or beneficiary associations, NGOs)

#### 3. Operational Capacity

◦ Technical expertise: The level of experience and skills in the prevention and control of STIs/HIV including genital chlamydia in young people aged 15-24 years.
◦ Geographic Coverage and Targets: The ability to reach all hard-to-reach regions or areas (remote, conflict) and target populations
◦ Monitoring and evaluation (M&E): the availability of annual reports and the ability to properly measure the impact of activities or interventions in a disaggregated manner.

### 8.2. Stakeholder Engagement Questionnaire for the Prevention and Control of Chlamydia in Young People Aged 15-24 in Mali and Burkina Faso

❖ Description of the structure and the person surveyed Q1. Countries: /.........../ Mali, Burkina Faso, Benin, Niger

Q2. Type of structure: Public /......./ Private /......../ Civil society organization /.........../

Q3. Structure Name: /.........../

Q4, Name of the Respondent:/.........../

Q5: Position of the Respondent in the Structure:/.........../

Q6: Number of years of work experience:/...... over 30 years........../

Q7: Number of years of work at the current position in the structure:/.........../

Q8: Gender of the Respondent: /.........../

Q9: Age range of the Respondent: /.........../ 1= < 35 years, 2= 35-45 years, 3= > 45 years

Q10. Have you served as a focal point:/.........../ 1= STI 2=HIV/AIDS 3=Gender 0= Not applicable

❖ **Questions for Stakeholder Identification and Categorization**

Q11. What are the interests of your structure for the prevention and control of STIs in general and chlamydia in particular among young people aged 15-24?................................................

Q12: What are your targets during your activities or interventions for the prevention and control of STIs in general and chlamydia in particular among young people aged 15-24? /.........../ 1+ school 2= Non-school 3= Academics 4=key population 5= Other vulnerable population (...........)

Q13. List 2-3 key activities of your structure for the prevention and control of STIs in general and chlamydia in particular in young people aged 15-24

**Activity 1**:............................................................................

◦ Occasion or time of year...........................................
◦ Targets.......................................................................................................................
◦ Number of participants (estimated)...........................................................................
◦ Budget..........................................................................................................................
◦ Main Source of Budget..............................................................................................

**Activity 2**:...................................................................................................................

◦ Occasion or time of year.................................................................................................
◦ Targets.........................................................................................
◦ Number of participants (estimated)............................................................................
◦ Budget.............................................................................
◦ Main Source of Budget.....................................................................................................

**Activity 3**:...................................................................................

◦ Occasion or time of year....................................................................................................
◦ Targets.............................................................................................................................
◦ Number of participants (estimated)..................................................................................
◦ Budget.............................................................................................................................
◦ Main Source of Budget...........................................................................................................

Q14. Who are the funders of your activities or interventions for the prevention and control of STIs in general and chlamydia in particular among young people aged 15-24? /............./

Landlord 1.....................................................................................................................................
Public /..................../ Private /.........../ CSO /............/ Private /.........../
Donor 2..........................................................................................................................................
Public /..................../ Private **/.........../** CSO **/............/** Private /.........../
Donor 3........................................................................................................................................
Public /..................../ Private /.........../ CSO /............/ Private /.........../

Q15. Where do you intervene or carry out your activities for the prevention and control of STIs in general and chlamydia in particular among young people aged 15-24?

◦ Country /.........../
◦ Region within the country/.........../
◦ City in the region /.........../
◦ Communes in the city /.........../

Q16. Are reports from the last 3-5 years available? YES /...../ No /.........../

Q17: How long have you been doing these activities or interventions for the prevention and control of STIs in general and chlamydia in particular in young people aged 15-24? /............./

1= less than 3 years, 2= 3-5 years, 3 = more than 5 years

Q18: In which development sector(s) do you intervene for the prevention and control of STIs in general and chlamydia in particular among young people aged 15-24?

1=health 2=agriculture 3=transport 4=urban planning 5=mining 6=security & uniforms 7=commerce 8=education 9=sport & youth 10=arts & tourism 11=other..............................

Q19. What is your structure’s short-, medium- and long-term plan for the prevention and control of STIs in general and chlamydia in particular among young people aged 15-24?

- Short-term (< 1 year)

◦ Objective...............................................................................................................................
◦ Human and Financial Resource Mobilization Plan..........................................
- Medium term (1-5 years)

◦ Objective...............................................................................................................................
◦ Human and Financial Resource Mobilization Plan..........................................
- Long term (> 5 years)

◦ Objective...............................................................................................................................
◦ Human and Financial Resource Mobilization Plan..........................................

Q20. If you are asked, how would you like to collaborate with the Pfizer FAPH chlamydia project in 2025?................................................................................................................

Q21. If asked, how would you like to collaborate with the Pfizer FAPH Chlamydia Project team beyond 2025 for the sustainability of this project?..................................

Q21. If you are asked, how would you like to collaborate with the Pfizer FAPH chlamydia project team beyond 2025 for the scale-up of this project?.....................................

Q22. Are you interested in taking part in the project restitution seminar in mid-December 2025? /........../ 1=Yes 2= No 3= I do not know

Q23. What is the best way to communicate with you? /.........../ 1= Phone 2= Email 3= In person by appointment 4=online (google meet or zoom 5= during a seminar 6. Other.....................

Q24. How often do you want to be informed about the progress and prospects of the project? /........../ 1= Monthly 2= Quarterly 3= Biannual 4 = Annual

Your whatsapp phone number:........................Your email.......................................

**Thank you for your participation**

